# Integrating bulk and single-cell RNA sequencing reveals *SH3D21* promotes hepatocellular carcinoma progression by activating the PI3K/AKT/mTOR pathway

**DOI:** 10.1101/2024.04.14.24305789

**Authors:** Wangxia Tong, Tao Lu, Li Liu, Rong Liu, Jibing Chen, Ning Luo

## Abstract

As a novel genetic biomarker, the potential role of *SH3D21* in hepatocellular carcinoma remains unclear. Here, we decipher the expression and function of *SH3D21* in human hepatocellular carcinoma. The expression level and clinical significance of *SH3D21* in hepatocellular carcinoma patients, the relationship between *SH3D21* and the features of tumor microenvironment (TME) and role of *SH3D21* in promoting hepatocellular carcinoma progression were analyzed based on the bulk samples obtained from The Cancer Genome Atlas (TCGA) and International Cancer Genome Consortium (ICGC) databases. Single-cell sequencing samples from Gene Expression Omnibus (GEO) database were employed to verify the prediction mechanism. Additionally, different biological effects of *SH3D21* on hepatocellular carcinoma cells were investigated by qRT-PCR, CCK-8 assay, colony forming assay and Western blot analysis. Bioinformatics analysis and *in vitro* experiments revealed that the expression level of *SH3D21* was up-regulated in hepatocellular carcinoma and correlated with the poor prognosis in hepatocellular carcinoma patients. *SH3D21* effectively promoted the proliferation, invasion, and migration as well as the formation of immunosuppressive microenvironment of hepatocellular carcinoma. In addition, *SH3D21* can activate the PI3K/AKT/mTOR signaling pathway. *SH3D21* stimulates the progression of hepatocellular carcinoma by activating the PI3K/AKT/mTOR signaling pathway, and *SH3D21* can serve as a prognostic biomarker and therapeutic target for hepatocellular carcinoma.

## 1. Introduction

Primary hepatoma is one of the most common malignant tumors, primarily hepatocellular carcinoma (HCC). It is often characterized by a high degree of malignancy, with a propensity for metastasis and recurrence following the treatment [1, 2]. Despite the availability of various treatment modalities, including surgical resection, radiotherapy, and chemotherapy [2, 3], the prognosis for HCC patients remains significantly poor [4]. Recent advancements in targeted therapy for HCC, such as application of vascular endothelial growth factor receptor (VEGFR) inhibitors and programmed cell death 1 ligand 1(PD-L1) inhibitors, have sparked newfound hope [5, 6]. However, the efficacy of targeted therapy is limited, thereby benefiting fewer than one-third of patients and only marginally prolonging the median survival by a mere three months [7]. Hence, identification of effective biomarker is urgently needed for targeted therapy of HCC.

The PI3K/Akt/mTOR pathway represents a crucial intracellular signaling cascade that can regulate diverse cellular processes including proliferation, apoptosis, metabolism, and angiogenesis, by functioning through intricate crosstalk with upstream or downstream molecules [8, 9]. Aberrant activation of the PI3K/Akt/mTOR pathway has been reported in approximately 50% of hepatoma cases [10], thereby exerting significant influence on the various facets of hepatoma initiation as well as progression, including tumor cell proliferation, differentiation, metabolism, autophagy, and immune regulation [11, 12]. Although multi-targeted tyrosine kinase inhibitors are being used as first-line therapeutic agents targeting the PI3K/Akt/mTOR pathway, their efficacy in improving patient survival remains modest. Notably, patients often develop acquired drug resistance during the treatment, thus limiting the therapeutic options available [13]. This could be possibly attributed to the limited understanding of the intricate interactions and metabolic roles of the PI3K/Akt/mTOR signaling axis in hepatoma [14].

SH3 domain-containing 21 (*SH3D21*) is a SH3 domain containing protein that is localized within the nucleus and plasma membrane. It functions by forming protein complexes and thereby modulating intracellular signaling to regulate various key biological processes, including cell division, differentiation, and growth and development [15]. Despite its biological significance, the current understanding of the role of *SH3D21* in human tumors remains limited. Given the dearth of experimental studies related to the role of *SH3D21* in tumorigenesis, this study was designed to elucidate its involvement in tumors and evaluate its potential utility both as a novel biomarker and therapeutic target.

The association between *SH3D21* and prognosis as well as clinical features in HCC patients were analyzed in this study. The mechanisms through which *SH3D21* can promote HCC and its effect on PI3K/Akt/mTOR pathway activation were also investigated. The clinical samples and cell experiments were employed to verify the experimental results.

## 2. Materials and methods

### Data source

The transcriptome and clinical characteristics analysis data of *SH3D21* in HCC were obtained from TCGA database (https://portal.gdc.cancer.gov) and ICGC database (https://dcc.icgc.org/)(download time: 20221224). We have also incorporated the genetic variation information obtained from the biological portal website (https://www.cbioportal.org/) [16]. Specifically, we have utilized a dataset comprising 442 liver samples obtained from TCGA and Firhose Legacy. The data of human pan-cancer was retrieved from the UCSC Xena online database (https://xena.ucsc.edu/) [17]. The downloaded data included mRNA expressions and the clinical features of 33 different tumors.

### Differentially expressed genes (DEGs) and analysis of characterization of tumor microenvironment

The “Limma” software package in the R language was used to identify the various differentially expressed genes (DEGs) and the expression levels of *SH3D21* in both the tumor tissues and control tissues in the dataset. The differential gene screening was identified using the criteria of log_2_FC>1.0 and FDR<0.05, thereby defining the genes exhibiting significant differential expression. We have stratified the tumor-derived samples into up- and down-regulation groups based on the median *SH3D21* expression level. The IOBR package in R language was employed to analyze the potential relationship between *SH3D21* expression level and the characteristics of tumor microenvironment by ssGSEA algorithm.

### GO (Gene Ontology), GSEA (Gene-set Enrichment Analysis) and KEGG (Kyoto Encyclopedia of Genes and Genomes) analyses of bulk samples

The tumor samples in TCGA data set were divided into up- and down-regulation groups based on the median expression level of *SH3D21* for the differential gene analysis. In addition, GO, KEGG and GSEA enrichment analysis of the differential genes were performed. The tumor samples in ICGC data set were also divided into up- and down-regulation groups according to the median expression level of *SH3D21* for differential gene analysis. Thereafter, the correlation between the expression level of *SH3D21* and the differential genes was analyzed (CoR=0.4, *P*<0.05). Subsequently, the high and low expression differential genes were fed into WebGestalt (www.WebGestalt.org), an online bioinformatics website that utilizes the KEGG database [18]. *P*<0.05 was considered to be a statistically different enrichment pathway.

### Single-cell sequencing sample quality control and cell type identification

The quality control process of the single-cell sequencing samples was conducted using Seurat (version 4.15) [19]. To ensure high-quality cells for the downstream analysis, the following criteria were applied: The cells expressing fewer than 500 or more than 6000 total genes were filtered out, cells with over 15% mitochondrial genes were excluded and the genes expressed in fewer than three cells were removed. Overall, a total of 60,437 cells were detected and the analysis incorporated 19,125 different genes. The batch effects were corrected using the Harmony R package, whereas the linear regression was employed to eliminate effects of the cell cycle. To identify the principal clusters, the Seurat FindClusters function (resolution = 0.5) was utilized and the top 30 principal component analysis (PCA) components were selected for further analysis using the gravel plot. The resulting clusters were thereafter visualized using “tSNE” and “UMAP” plots. The FindAllMarkers function of Seurat was employed to identify cell markers for each cluster. To identify major cell types and provide annotations, the CellMarker database (http://xteam.xbio.top) was combined with the data source of the cell surface markers [20].

### InferCNV

The R software copyKAT package was used for CNV analysis of the hepatocytes present in the samples, and the T cells were used as normal reference cells. The normal hepatocytes and HCC cells were distinguished according to the changes of CNV.

### Differential gene identification and GSVA (Gene-set Variation Analysis) enrichment analysis in the single-cell samples

A nonparametric and unsupervised algorithm for gene set variation analysis (GSVA) was utilized to evaluate the pathway activity in normal hepatocytes, *SH3D21*-positive HCC cells, and *SH3D21*-negative HCC cells. The geneset used for GSVA enrichment analysis was Hallmark geneset.

### Pseudotime trajectory analysis

The Monocle2 package (v2.8.0) of R software was employed to perform the pseudo-time series analysis of HCC cells [21]. The input for the analysis consisted of the original count matrix obtained from the data processed by Seurat. The new Cell Data Set function was utilized to create an expression family object, facilitating the specification of the lower detection limit as 0.5. An unsupervised analysis method was employed to explore the cell development trajectory using the highly variable genes selected by Monocle. The dispersion empirical parameter was set to 1, whereas the remaining data analysis parameters were selected as the default settings.

### Immunotherapy prediction and chemotherapy sensitivity analysis

Three distinct immunotherapy cohort studies GSE78220, GSE67501 and IMvigor210 were collected from GEO database to examine the potential correlation between *SH3D21* and immunotherapy. The tumor immune atlas database (The Cancer Immunome Atlas, https://tcia.at/)was also used to observe the treatment response of *SH3D21* to PD1 and CTLA4. In addition, based on Cancer Drug Sensitivity Genomics (GDSC), a public pharmacology portal, the chemotherapeutic agents sensitive to *SH3D21* were estimated and the top 10 drugs were shortlisted for further analysis.

### Test and verify

#### Cell lines and cell culture

LX-2, Huh7, HepG2, SK-HEP-1 cell line was provided by CTCC, Ltd (Wuxi, China). The frozen HepG2 (in Dulbecco’s modified Eagle medium (DMEM, high glucose)), Huh-7 (in DMEM, high glucose, and SK-HEP-1 (MEM-EBSS) cells were thawed at 37 ℃ and then resuspended into the corresponding complete culture media. When the cells grew to approximately 80%, cell passage was executed. Additionally, the cells were grouped and cultured in an incubator (Thermo, USA) at 37 ℃. The cells were thereafter collected and then stored for subsequent experiments.

#### Tissue source

The HCC tissues and normal hepatic tissue were procured from the different individuals undergoing liver cancer resection or liver biopsy at the Department of Hepatobiliary Surgery, Ruikang Hospital Affiliated to Guangxi University of Traditional Chinese Medicine between December 2022 and October 2023.The studies were approved by the Ethics Committees of Ruikang Hospital Affiliated to Guangxi University of Traditional Chinese Medicine. The assigned ethical review approval number: KY2022-045.

#### Real-time, quantitative reverse transcription-polymerase chain reaction (qRT-PCR)

Total RNA was extracted by using Trizol reagent (Invitrogen, Grand Island, New York, USA). Both the concentration and purity of RNA were measured using a Nanodrop spectrophotometer (ThermoFisher Science, Waltham, Massachusetts, USA). RNA was reverse transcribed into cDNA using a reverse transcription kit (Takara, Dalian, China) in accordance with the manufacturer’s instructions. SYBR Select Master Mix (ThermoFisher Science, Waltham, Massachusetts, USA) was used for qRT-PCR analysis. A QuantStudio^TM^ 6 Flex real-time quantitative PCR system was employed for the data collection. The samples obtained amplified in accordance with the following protocol: 95°C 5 mins, 95°C 40 cycles 30s, 60°C 40s, 72°C 1 min. Glyceraldehyde-3-phosphate dehydrogenase (GAPDH) was used as an endogenous control. Each sample was prepared in triplicate. The 2^−ΔΔCT^ method was used to calculate the relative expression. The sequences of the qRT-PCR primers used for *SH3D21* were: forward: 5’-AGCAAGGAGGGCAATGACTCT-3’, reverse: 5’-ACGCAGTACTTGCCACTCT-3’; GAPDH: forward: 5’-GCAGCGTGATCCCTGCAAAAT-3’, reverse: 5’-GCCTGTAGTCAGCAACTCATCG-3’.

#### Cell transfection

Huh7 and HepG2 cells were seeded in six-well culture plate for 24 h (1×10**^5^** cells/well). Thereafter, the cells were divided into the HepG2 and Huh7 group, the HepG2/Huh7+siRNA NC group and the HepG2/Huh7+si-*SH3D21*-1, 2, 3 group. HepG2/Huh7+si-*SH3D21*-1, which was validated by RT-PCR to be the optimized knockdown target sequence, was selected for the subsequent experiments. The DNA sequence of *SH3D21* gene was amplified by PCR, digested, and then cloned into the expression vector (PCI-neo). The expression vector was then transfected into HepG2 cells to enable up-expression of the *SH3D21* gene; the group was confirmed to be the OE-*SH3D21* if the up-expression was validated by RT-PCR. Additionally, Lipofectamine 2000 transfection reagent (1.5 μL/mL, ThermoFisher, USA) was used to transfect 50 nM gene fragments or 1 μg of plasmid to HepG2 and Huh7 cells. After 48-hour transfection, the samples were collected and stored.

#### Colony formation

The samples were divided into the control group, the si-control group and the si-*SH3D21* group. After transfection, the cell suspension was prepared once the cell growth was in the logarithmic phase. The cells were then seeded into the six-well plates (Corning, USA) and incubated at 37 ℃, 5% CO_2_ and saturated humidity for two weeks. The culture was terminated if the clones visible to the naked eyes appeared in the dish. Afterward, the cells were rinsed with PBS twice and fixed with 5 mL of pure methanol (Beijing Dingguo Changsheng Biotechnology Co., Ltd., China) for 15 min. The methanol was then removed, and Giemsa staining solution (Beyotime, China) was added and incubated for 30 min. The staining solution was removed, and the sample was dried in air. The cells were counted manually from obtained images. Alternatively, clones with more than 10 cells were counted using a light microscope (Olympus, Japan). The colony formation rate was calculated by using the formula (number of clones)/(number of inoculated cells) × 100%.

#### Determination of cell viability

Cell Counting Kit-8 (CCK-8) cell proliferation and cytotoxicity assay kit (Sigma-Aldrich, Merck KGaA, Germany) was used to measure the viability of transfected Huh7 and HepG2 cells. The cells were seeded in the wells of a 96-well plate and thereafter incubated for 24h, 48h, 72h, respectively. CCK-8 solution (15 µL) was added to each well at the designated time intervals and incubated at 37°C for 4h. The absorbance at 492 nm was measured using a microplate reader (Thermo Scientific, Waltham, Massachusetts, USA). Each experiment was repeated independently in triplicate.

#### Western blot analysis

HCC cells and tissues were lysed in radioimmunoprecipitation assay (RIPA) buffer (KeyGEN, Nanjing, China) containing protease suppressors for 30 min. The protein concentration was quantified using a bicinchoninic acid (BCA) kit (KeyGEN, Nanjing, China) in accordance with the manufacturer’s protocol. The total protein was subjected to heating at 95°C for 5 min. Thereafter, identical quantities of protein were separated by sodium dodecyl sulfate polyacrylamide gel electrophoresis (SDS-PAGE). The separated proteins were transferred to polyvinylidene fluoride (PVDF) membranes after which electrometastasis was applied and then blocked with 5% skimmed milk at the room temperature for 60 min. The membranes were then stored overnight at 4°C with an appropriate primary antibody. The panel of antibodies used was as follows: *SH3D21*(1:1000; Proteintech No. 25767-1-AP, Chicago, USA), mTOR (1:1000; Proteintech No. 66888-1-lg, Chicago, USA), p-AKT (1:1000; Proteintech No. 66444-1-lg, Chicago, USA), p-PI3K (1:1000; Proteintech No. 4228, Chicago, USA), PI3K (1:1000; Proteintech No. 20584-1-AP, Chicago, USA), AKT (1:4000; Proteintech No. 60203-2-lg, Chicago, USA), and GAPDH (1:5000; Proteintech No. 60004-1-Ig, Chicago, USA). After washing 3 times in TBST buffer, the membranes were incubated at the room temperature for 1h with rabbit horseradish peroxidase (HRP)-conjugated secondary antibody (1:1000, Beyotime. No. A0208, Shanghai, China) then washed with the blocking solution and visualized by enhanced chemiluminescence (ECL, Thermo Fisher Scientific, Waltham, Massachusetts, USA). Finally, quantity One gel analysis software was used to detect the signal intensity of each membrane. The intensity was measured relative to that of GAPDH.

#### Immunohistochemical staining

Following the dewaxing and antigen retrieval steps, the tissue sections were subjected to additional treatments. First, incubation with 3% H2O2 (Beyotime, China) for 10 min and 0.1% Triton X-100 (Beyotime, China) at the room temperature for 10 min was conducted. Subsequently, 50 μL of 5% BSA (Wuxi Puhe, China) was applied to the tissues and incubated at 37 ℃ for 1 hour to block non-specific binding sites. The primary antibody, specifically SH3D21 (dilution: 1:1000; Proteintech No. 25767-1-AP, Chicago, USA), was proportionally diluted and then applied to the tissues, followed by incubation at 37 ℃ for 2 h. After washing with PBS, a secondary antibody was added: Rabbit secondary antibody (dilution: 1:200, Zhongshan Jinqiao, China) or Murine dimab (dilution: 1:200, Zhongshanjinqiao, China), depending on the species sources of the primary antibodies used. Thereafter, incubation with the secondary antibody was performed at 37℃ for 1h. For visualization, diaminobenzidine (DAB) (Nakasugi Kinbashi, China) color development was employed. Hematoxylin restaining was then conducted, followed by sealing with the neutral gum. Finally, the prepared tissue sections were observed and photographed under a microscope (Olympus, Japan) to evaluate the target staining and overall tissue morphology.

#### Invasion and Migration Experiments

Matrigel matrix glue (BD Biosciences, USA) was diluted in a serum-free medium at 1:6, and 50ul was evenly spread into the upper chamber of Transwell chamber (Corning, USA). The chamber was placed in a 24-wellplateandincubated at 37°C for 4h to allow it to gelatinize. The cell density was adjusted to 2×105 cells/mL, and 100μL per well was inoculated into the upper chamber of transwell chamber. The cells in the upper chamber were removed after 24h, fixed with methanol and 0.1% crystal violet (Beijing Dingguo Changsheng Biotechnology, CHN) for 20min, stained for 10min, and washed twice with PBS. The cells were then counted under an inverted optical microscope (Olympus, JPN).

#### Scratch Wound-Healing Assay

Huh7 and HepG2 cells were seeded in 6-well culture plates (Corning, USA) at 95–100% confluence. The cells were harvested 48h after transfection. The scratch was created using a 200 µL pipette tip. PBS was used to wash the plates thrice to remove the cellular debris. The images for the wound closure were captured at the different times (12, 24, 36 and 48 h) by microscope (Olympus, JPN).

### Statistical analysis

The biological information analysis in this study was conducted by using R programming (version 4.15, https://www.r-project.org/). Statistical analysis was performed by SPSS 27 and GraphPad Prism V7.0 software. The data has been presented as mean ± standard deviation (SD). For comparison of means between the two groups, T-test was applied. Analysis of variance (ANOVA) was employed to compare multiple sets of values. The relationship between the expression of *SH3D21* and clinical features was assessed using the Chi-square test. Survival analysis was performed using the Kaplan-Meier method, and the log-rank test was used for the group comparisons. *P*<0.05 was considered statistically significant.

## 3. Results

### *SH3D21* was highly expressed in HCC and related to poor outcomes of HCC patients

The differential analysis of HCC transcriptome data from TCGA database (https://portal.gdc.cancer.gov) revealed that the mRNA expression of *SH3D21* in HCC tissued was significantly higher than that in normal hepatic tissue (logFC=1.937) (**Fig 1A**). Moreover, determination of the relationship between *SH3D21* expression and clinical characteristics of HCC patients revealed that *SH3D21* was highly expressed in females (*P*=0.0099) and increased with the elevation of tumor T-stage (**Fig 1B**), histological grading (**Fig 1C**) and clinical stage (**Fig 1D**). High expression of *SH3D21* resulted in shorter survival of HCC patients (**Fig 1E**). However, no correlation was observed between Progression Free Survival (PFS), Disease Free Interval (DFI), and Disease-Specific Survival (DSS) (*P*>0.05). Univariate and multivariate analyses were conducted to investigate the various clinical features and the possible relationship between *SH3D21* expression and the survival status of patients. High expression of *SH3D21* remained a significant independent predictor for patients with HCC (**Fig 1I, J**). In addition, HCC dataset (LIRI-JP) was downloaded for validation from the ICGC (https://dcc.icgc.org/) database. The results revealed that *SH3D21* was highly expressed in HCC (logFC= 1.442) (**Fig 1F**) and increased with both the elevated histological grading (**Fig 1G**) and clinical stage of tumor tissues (**Fig 1H**). The above results illustrated that *SH3D21* was highly expressed in HCC and was associated with poor clinical outcomes in HCC patients. To experimentally validate the predicted results, three HCC-derived samples and normal liver tissue-derived samples were collected. Immunohistochemical staining and Western blot analysis to determine the expression of *SH3D21* protein in HCC and normal liver tissues was performed. The results demonstrated that the level of *SH3D21* protein was significantly higher in the tumor samples compared to the normal liver tissues (**Fig 1K-M**) **(Full length gel Supplementary Fig 1)**. These results confirmed the prediction that the expression of *SH3D21* was increased in HCC.

**Fig: 1.**
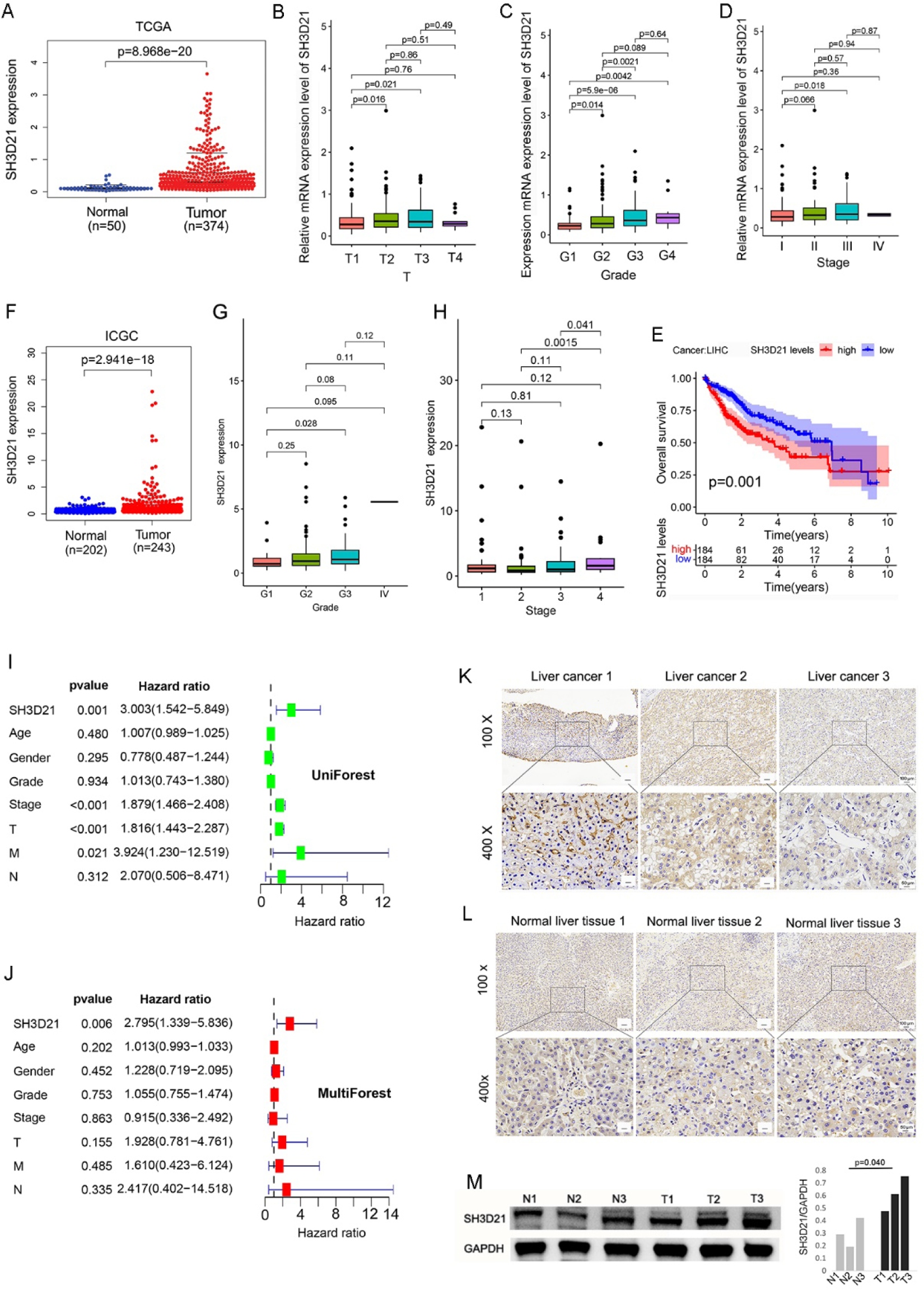
*SH3D21* was highly expressed and associated with poor clinical outcomes in HCC patients. **A** Differential analysis of mRNA expression of *SH3D21* in normal control and HCC tissues obtained from TCGA database source samples. **B-D** Relationship between the expression level of *SH3D21* and T Stage (B), Grade (C), and stage (D) of HCC patients in TCGA database. **E** Impact of high expression of *SH3D21* on prognosis of HCC patients. **F** Differential analysis of mRNA expression of *SH3D21* in the normal control and HCC tissues from ICGC database source samples. **G, H** Correlation between *SH3D21* expression level and Grade (G) and Stage (H) in ICGC database source samples. **I, J** Univariate and multifactorial analysis of the clinical characteristics of HCC patients and correlation between *SH3D21* gene expression and survival status of patients from TCGA database. **K, M** The expression of *SH3D21* protein in HCC and normal liver tissue samples was detected by immunohistochemical staining and Western blotting.

### Determination of Mutation as well as methylation status of *SH3D21* in HCC and relationship between *SH3D21* expression and TME

To investigate the impact of *SH3D21* genetic variation on mRNA expression, the mutation of *SH3D21* in HCC samples was next analyzed using the online database ccBioPortal (https://www.cbioportal.org/). As demonstrated, our analysis revealed that *SH3D21* was genetically altered in 6.88% (24/349) of the samples. Among these alterations, the highest frequency was observed in cases of high mRNA expression (20/349, 5.73%), followed by mutations (2/349, 0.57%), amplification, and deep deletion (1/349, 0.29%) (**Fig 2A**). The most common mutation in the *SH3D21* gene was the I428T missense mutation (0.07%), which was diploid in nature. The second most frequent mutation was the M355Cfs*38 frameshift mutation (0.06%), resulting in a mild copy number loss (**Fig 2B**). Further correlation analysis between the mutation type and mRNA expression of *SH3D21* revealed that the increase in mRNA expression could be primarily attributed to gene expansion and significant amplification events (**Fig 2C**). Therefore, mRNA expression of *SH3D21* was negatively related to the methylation level **(Fig 2D).** Overall, these findings indicated that *SH3D21* mRNA expression levels were associated with gene amplification.

**Fig. 2.**
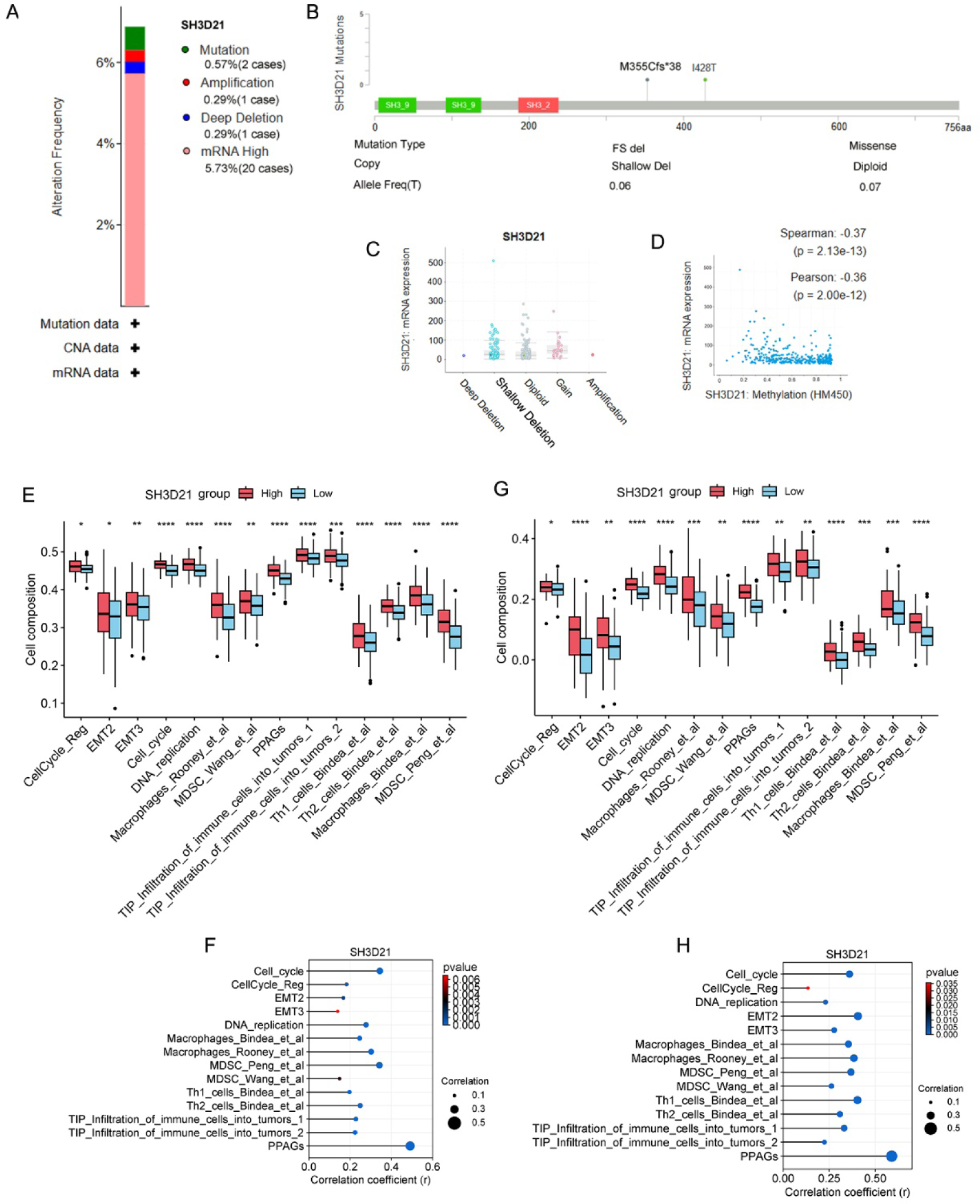
Mutation and methylation analysis of *SH3D21* in HCC and the relationship between *SH3D21* expression and TME features. **A** The type and frequency of the genetic changes in *SH3D21*. **B** *SH3D21* mutation type, site, and proportion. **C** Potential relationship between *SH3D21* mRNA expression and mutation. **D** The relationship between *SH3D21* expression and methylation. **E** Relationship between *SH3D21* gene expression and TME characteristics in the TCGA dataset. **F** Correlation analysis between *SH3D21* gene expression and TME characteristics in TCGA dataset. **G** Relationship between *SH3D21* gene expression and TME characteristics in the ICGC dataset. **H** Correlation analysis between *SH3D21* gene expression and TME characteristics in ICGC dataset.

To explore the possible function of *SH3D21* in promoting the clinical progression of HCC, TCGA and ICGC datasets were employed to analyze the correlation between the expression of *SH3D21* and the microenvironment characteristics of HCC. The results revealed that increase of *SH3D21* expression was associated with EMT2, EMT3, cell cycle, DNA replication, TH1/TH2 cell regulation, MDSC and TIP tumor immune cell infiltration. Therefore, we speculated that there was a significant correlation between the above characteristics and the expression level of *SH3D21*. In addition, the above characteristics displayed a consistent trend in TCGA **(Fig 2E, F)** and ICGC data sets **(Fig 2G, H)**, thereby indicating that *SH3D21* can play a vital role in regulating the proliferation, invasion, migration, and formation of immunosuppressive microenvironment of HCC.

### *SH3D21* can promote the proliferation, invasion, and migration of HCC cells as confirmed by *in vitro* experiments

Three distinct kinds of HCC cells, SK-HEP-1, Huh-7 and HepG2 were selected to observe the mRNA and protein expression of *SH3D21*. The results revealed that the mRNA and protein expression levels of *SH3D21* were markedly higher in the three HCC cells in comparison to the control LX-2 cells (**Fig 3A, B**) **(Full length gel Supplementary Fig 2)**. Interestingly, silencing of *SH3D21* in HepG2 and Huh-7 cells resulted in reduced cell viability at 48h and 72h (*P*<0.01) (**Fig 3C, F**) and a significant decrease in the cell colony forming ability (*P*<0.01) (**Fig 3D, E**). Moreover, silencing the expression of *SH3D21* significantly inhibited the wound healing ability (**Fig 3G, I**), as well as reduced the invasive ability (**Fig 3H, K**) and the number of migrating HCC cells (**Fig 3H, J**). The above results confirmed that high expression of *SH3D21* can promote the proliferation, migration, and invasion of HCC cells.

**Fig. 3.**
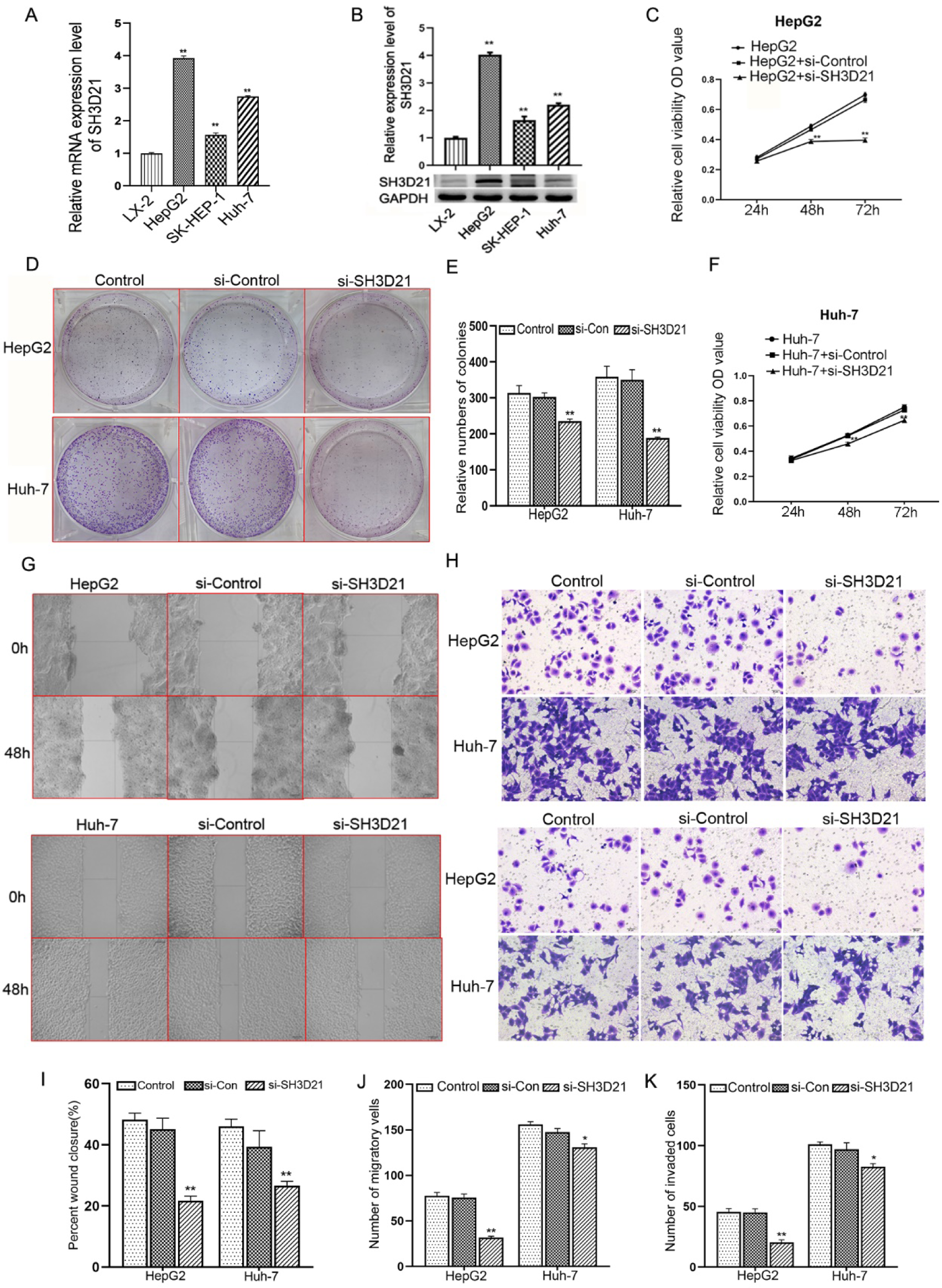
*SH3D21* was highly expressed and promoted proliferation, invasion, and migration of HCC cells. **A, B** *SH3D21* exhibited higher mRNA and protein expression levels in HCC cell lines than in human hepatic stellate cells. **C, F** Silencing of *SH3D21* gene significantly reduced the survival of HepG2 and Huh-7 cells at 48h and 72h, respectively. **D, E** Silencing *SH3D21* markedly inhibited colony-forming ability of HCC cells. **G, I** Silencing *SH3D21* significantly suppressed wound healing ability of HCC cells. **H, K** Silencing *SH3D2* significantly reduced the invasive ability of HCC cells. **H, J** Silencing *SH3D2* significantly attenuated the number of migrating HCC cells.

### Bulk sample analysis of TCGA and ICGC databases revealed that *SH3D21* can cause activation of PI3K/AKT signaling pathway in HCC

HCC samples in the TCGA dataset were divided into high- and low-expression groups according to the median value of *SH3D21* expression for the differential analysis, as well as GO **(Fig 4A)** and KEGG enrichment analysis **(Fig 4B)** were performed for the differential genes. Biological process (BP) results of GO enrichment analysis revealed that *SH3D21* was mainly involved in organic acid metabolic process, carboxylic acid metabolic process and ion transport. Cellular component (CC) results demonstrated that *SH3D21* was located on the cell surface, intrinsic component of plasma membrane and integral component of plasma membrane. Molecular function (MF) of *SH3D21* included transmembrane transporter activity, oxidoreductase activity and transporter activity. KEGG enrichment analysis revealed that the differential genes were mainly enriched in regulation of retinol metabolism, metabolism of xenobiotics by cytochrome P450 and metabolic pathways. In addition, HCC samples in the TCGA dataset were divided into high- and low-expression groups according to the median value of *SH3D21* expression for GSEA pathway enrichment analysis. GSEA enrichment analysis further indicated that the differential genes associated with *SH3D21* high-expression group were enriched in cell cycle, DNA replication, mismatch repair, tight linking and PI3K-AKT-mTOR pathway. All these pathways are primarily related to HCC cell proliferation and invasion. The differential genes associated with *SH3D21* low-expression group were found to be enriched in drug metabolism cytochrome P450, fatty acid metabolism, complement and coagulation cascades and PPAR signaling pathway **(Fig 4C)**.

**Fig. 4.**
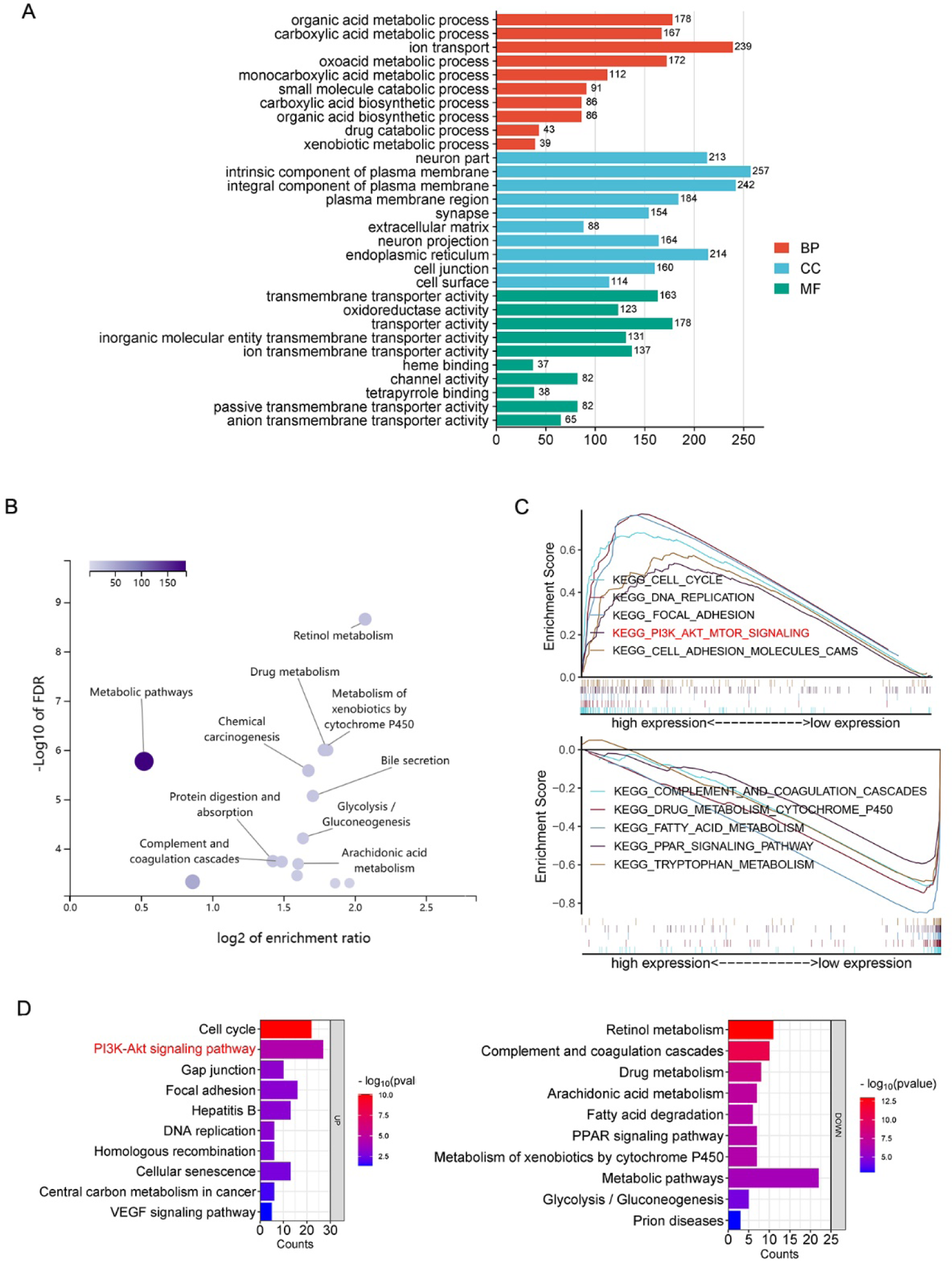
*SH3D21* promoted substantial activation of PI3K/AKT signaling pathway in HCC. **A, B** GO and KEGG enrichment analysis of differential genes associated with *SH3D21* high and low expression groups in TCGA dataset. **C** GSEA enrichment analysis of differential genes associated with *SH3D21* high and low expression groups in TCGA dataset. **D** KEGG enrichment analysis of differential genes associated with SH3D21 high and low expression groups in ICGC dataset.

KEGG enrichment analysis of *SH3D21-*associated differential genes was performed using the ICGC dataset to validate GSEA results from the TCGA dataset. The results suggested that the differential genes associated with *SH3D21* high-expression group were mainly enriched in pathways related to the tumor proliferation and invasion, such as cell cycle, PI3K-AKT signaling pathway, DNA replication, mismatch repair, gap linking, and VEGF angiogenesis pathway. The differential genes associated with *SH3D21* low-expression group were enriched in complement and coagulation cascades, drug metabolism, arachidonic acid metabolism and PPAR signaling pathway (**Fig 4D**). The above results were similar to the GSEA enrichment analysis results of TCGA dataset. Among the pathways enriched by the differential genes in *SH3D21* high-expression group, PI3K-AKT signaling pathway was identified. The PI3K-AKT signaling pathway can promote multiple signaling processes and regulate a variety of cellular functions, such as metabolism, proliferation, cell survival, growth, and angiogenesis. Activation of PI3K pathway can not only promote cell cycle progression, migration, but also inhibit cellular inflammation and apoptosis. Therefore, the above results indicated that the high expression of *SH3D21* has a potential role in activating PI3K signaling cascade.

### Single-cell sequencing analysis and *in vitro* experiments revealed that *SH3D21* can activate PI3K/AKT signaling pathway in HCC cells

Following data acquisition from Lu *et al.* [22], which involved single-cell sequencing of hepatoma from the GEO database, we focused on analyzing the single-cell sequencing results obtained from 10 different HCC tissues and 8 para-cancerous control tissues in the dataset. After the batch correction was performed using the harmony algorithm to eliminate its potential effect on the cell clustering **(Fig 5B)**. This resulted in the formation of distinct 18 cell groups **(Fig 5A)**. We utilized marker genes provided in the source literature to perform the cell type annotation **(Fig 5C)**. Subsequently, HCC cells, macrophages, monocytes, B cells, T cells and NK cells were selected for further analysis **(Fig 5D)**. Notably, our cell type annotation revealed that *SH3D21* was expressed in all the above 6 cell type **(Fig 5E)**.

**Fig. 5.**
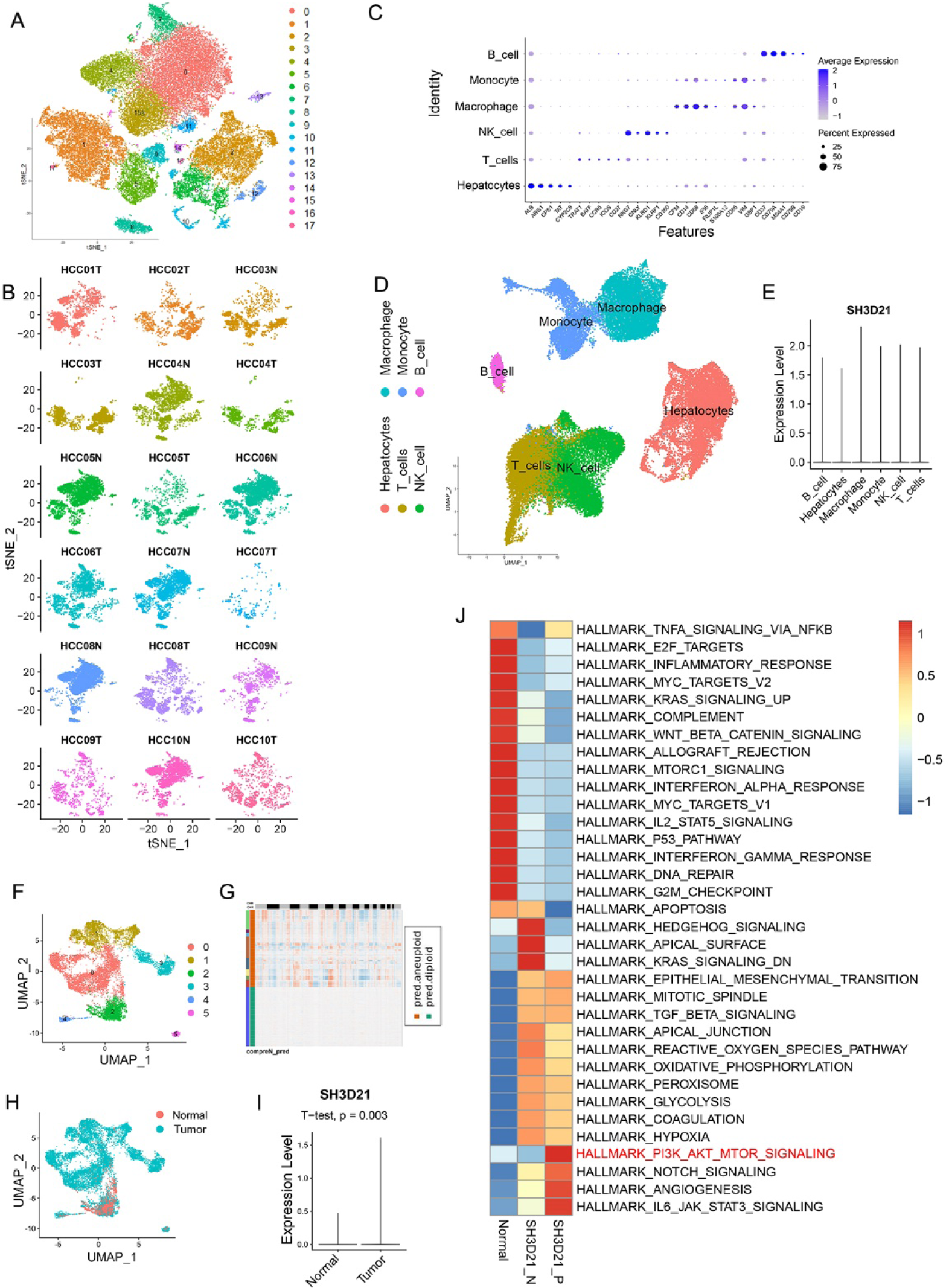
Single-cell sequencing revealed that *SH3D21* caused activation of PI3K/AKT signaling pathway in HCC. **A** For clustering of cells after the batch correction. **B** For clustering of cells after the batch correction was performed using the harmony algorithm. **C** Expression of the various cell marker genes in clustering cells. **D, E** Cell types that were included in subsequent analyses and expression of *SH3D21* in various cell types. **F-H** For clustering of subsets of hepatocytes(F), Copykat was used to perform the cellular CNV analysis on hepatocytes(G). Distribution of the normal hepatocytes and HCC cells (H). **I** Difference analysis of *SH3D21* expression between hepatocytes and HCC cells. **J** GSVA enrichment analysis of *SH3D21*-positive, *SH3D21*-negative HCC cells and the control hepatocytes.

The hepatocyte subsets were further analyzed, and cellular CNV analysis was performed on hepatocytes using Copykat **(Fig 5F).** After the diploid and heteroploidy of hepatocytes were identified, normal hepatocytes and HCC cells were distinguished **(Figs 5F-H)**. The differential analysis between the hepatocytes and HCC cells revealed that *SH3D21* was highly expressed in HCC cells **(Fig 5I)**. The hepatocyte subsets were further divided into *SH3D21* positive HCC cells, *SH3D21* negative HCC cells and normal hepatocytes, and GSVA enrichment analysis was conducted on the above three groups. The results of GSVA enrichment analysis revealed that *SH3D21* positive HCC cells exhibited enrichment in the different pathways associated with tumor proliferation, invasion, and immunosuppression, such as PI3K/AKT/mTOR, angiogenesis, EMT, IL6-JAK-STAT3. Interestingly, *SH3D21* negative HCC cells exhibited enrichment in various pathways associated with hedgehog signaling, apical junction, coagulation, and hypoxia. GSVA enrichment analysis revealed that normal hepatocytes exhibited enrichment in various pathways associated with inflammation and immune response, such as TNFA signaling via NFKB, inflammatory response, complement, P53, apoptosis **(Fig 5J).** These results indicated that *SH3D21* was highly expressed in HCC cells and can potentially activate the PI3K/AKT/mTOR pathway, thus promoting the growth of HCC.

Furthermore, *in vitro* experiments to examine above results. we observed that overexpression of *SH3D21* led to increased protein levels of mTOR, p-PI3K/PI3K, and p-AKT/AKT in HepG2 and Huh-7 cells **(Figs 6A, B) (Full length gel Supplementary Fig 3).** Conversely, when *SH3D21* was knocked down, reverse effects on the aforementioned proteins in HepG2 and Huh-7 cells were observed **(Figs 6C, D) (Full length gel Supplementary Fig 4)**. These findings indicated that *SH3D21* could activate PI3K/AKT/ mTOR signaling pathway in HCC cells.

**Fig. 6.**
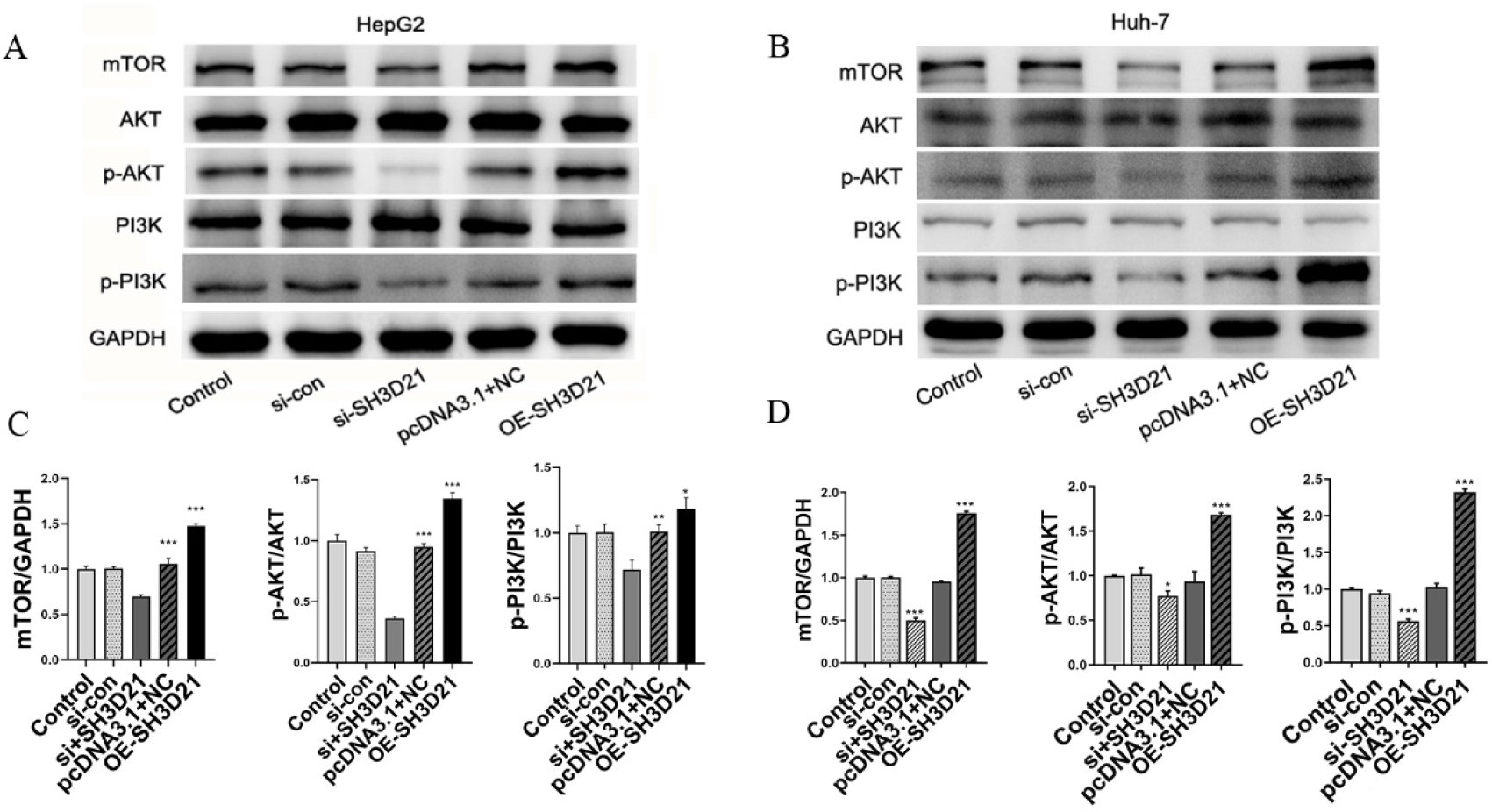
*In vitro* experiments revealed that *SH3D21* can activate the PI3K/AKT signaling pathway. **A, B** Protein expression electrophoretograms of key proteins in PI3K/AKT signaling pathway after *SH3D21* silencing and overexpression in HepG2 and Huh-7 cells, respectively. **C, D** Ratio of the gray value of key proteins in PI3K/AKT signaling pathway after *SH3D21* silencing and overexpression in HepG2 and Huh-7 cells.

### Immune landscape of *SH3D21* in HCC and its response to immunotherapy

The correlation analysis between *SH3D21* expression and TME based on TCGA database revealed that high expression group of *SH3D21* possessed higher immune score than low expression group of *SH3D21* (*P*<0.001)**(Fig 7A)**. This result is also confirmed in ICGC database analysis **(Fig 7B)**. However, the analysis results of TCGA and ICGC databases revealed that there was no statistically significant correlation observed between the expression level of *SH3D21* and immune scores (*P*>0.05). In addition, TCGA and ICGC datasets also revealed that there was no statistical difference observed in tumor matrix score and tumor purity score between high- and low-expression groups of *SH3D21*. The CIBERSORT method was employed to analyze the possible relationship between expression of *SH3D21* and immune cell infiltration in TCGA data set. The results revealed that high expression of *SH3D21* accompanied by M0 immune cell infiltration (*P*<0.001) **(Fig 7C)**. Moreover, the high expression of *SH3D21* was positively correlated with M0 cell infiltration (R=0.5, *P*=7.1E-08) **(Fig 7D).** This result was also confirmed in ICGC database analysis **(Fig 7E).** Based on analysis results of the immune microenvironment characteristics before, it was suggested that *SH3D21* could regulate immunity through affecting macrophages, MDSC and Th2 cells.

**Fig. 7.**
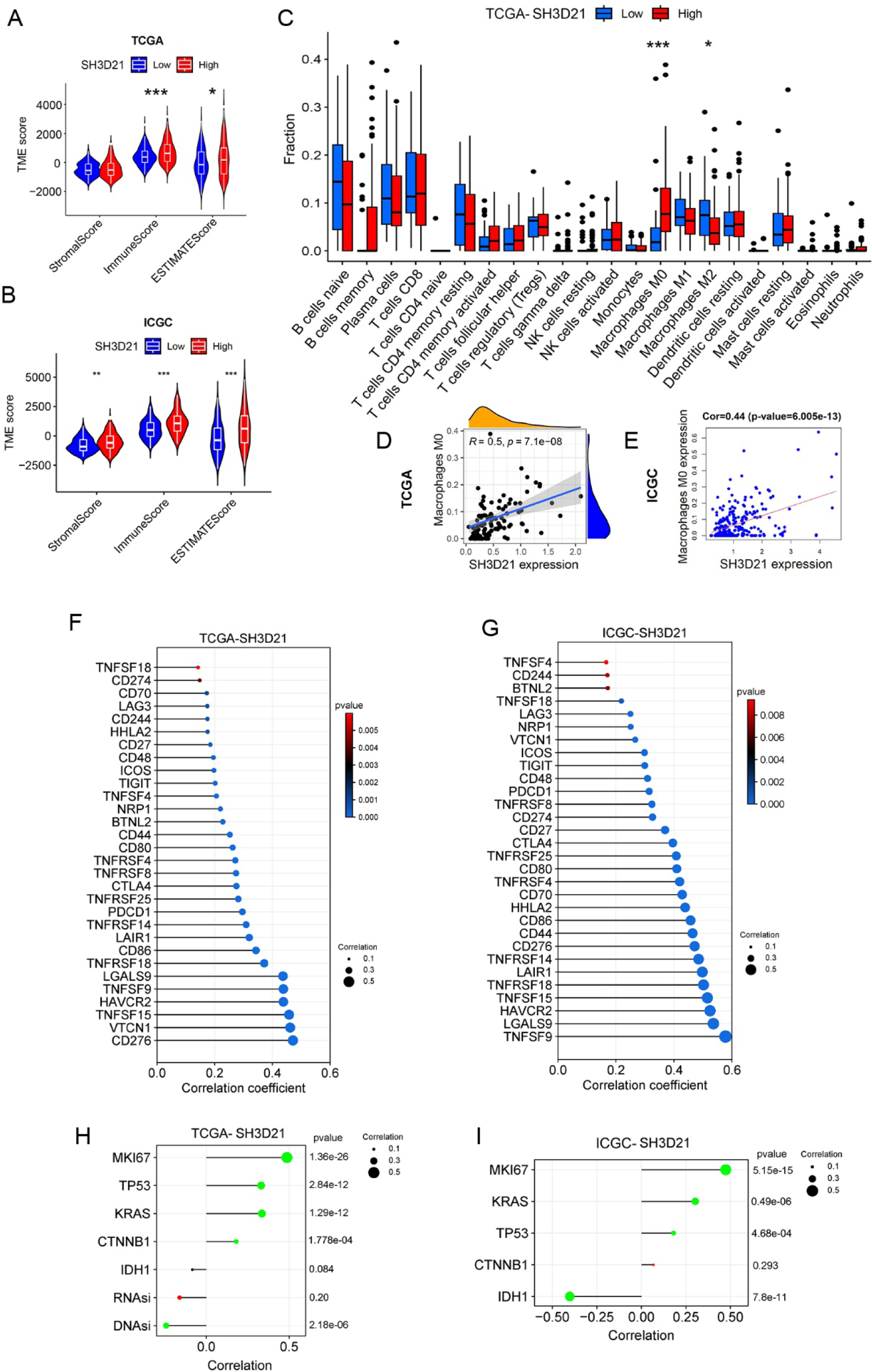
Immune landscape of *SH3D21* in HCC. **A, B** The correlation between expression of *SH3D21* and TME based on TCGA and ICGC data sets. **C** The correlation between high- and low-expression of *SH3D21* and immune cell infiltration. **D, E** The correlation analysis of expression of *SH3D21* and M0 cell infiltration based on TCGA and ICGC data sets. **F, G** The correlation analysis of high-expression of *SH3D21* and expression of immune checkpoint molecular based on ICGC and TCGA datasets. **H** The correlation analysis between *SH3D21* and MIK67, TP53, KRAS, CTNNB1and IDH1, and correlation analysis between *SH3D21* and DNAsi and RNAsi based on the TCGA dataset. **I** The correlation analysis between *SH3D21* and MIK67, TP53, KRAS, CTNNB1and IDH1 based on ICGC dataset.

**Fig. 8.**
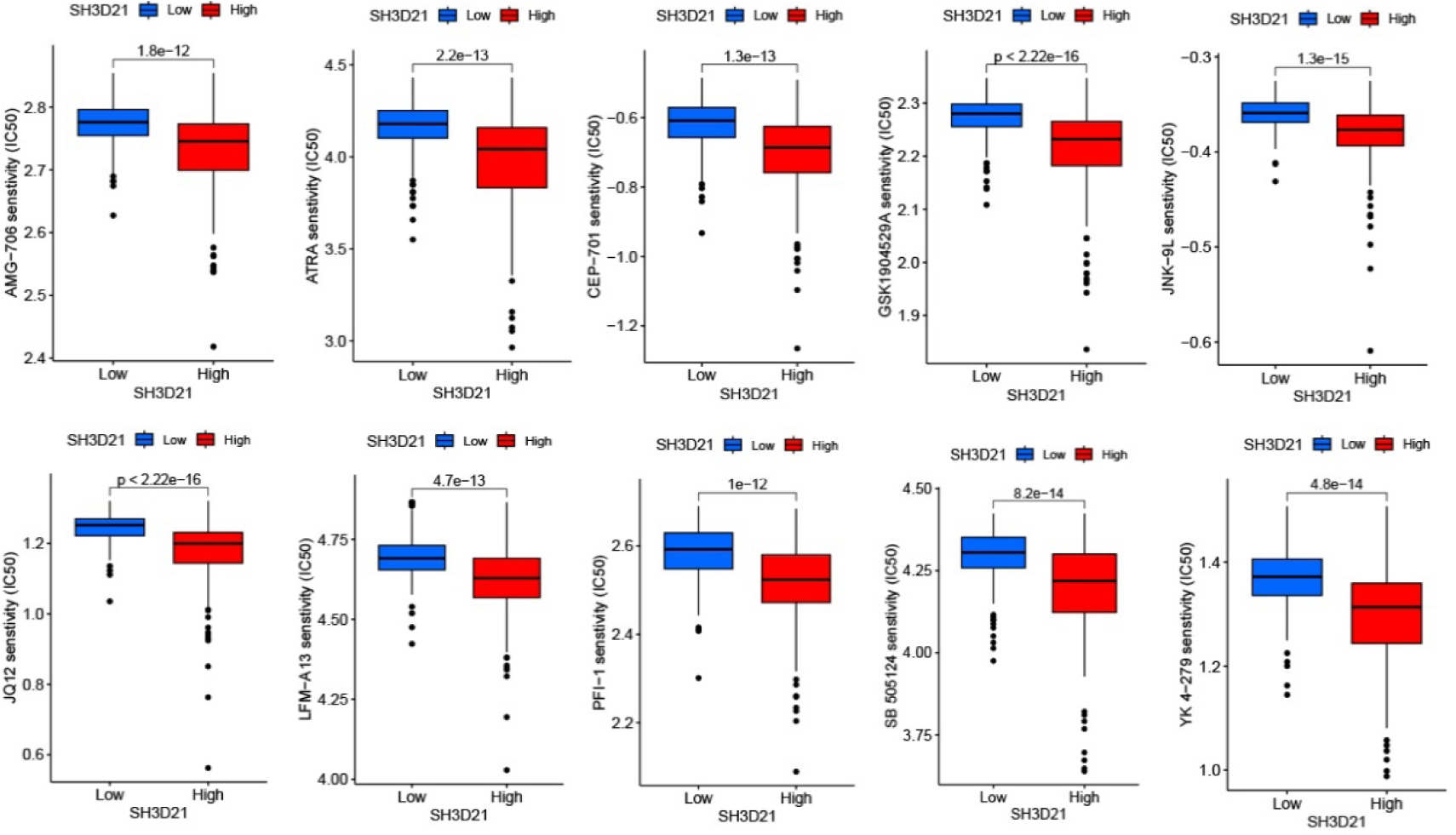
Top 10 sensitive drugs to *SH3D21* in drug sensitivity experiments.

The correlation analysis between *SH3D21* and immune checkpoints in TCGA **(Fig 7G)** and ICGC **(Fig 7F)** datasets demonstrated that high expression of *SH3D21* was positively correlated with the expression of immune checkpoint molecule. This observation indicated that high expression of *SH3D21* can affect immunosuppression microenvironment of HCC. In addition, the correlation between *SH3D21* and the expression level of tumor-related factors MKI67, TP53, KRAS, CTNNB1 and IDH1 were also investigated. The analysis based on TCGA **(Fig 7H)** and ICGC **(Fig 7I)** datasets revealed that the expression of *SH3D21* was positively correlated with MIK67, TP53, KRAS as well as CTNNB1, and the expression of *SH3D21* was negatively correlated with IDH1. In addition, the correlation analysis between *SH3D21* and the stemness of HCC cells revealed that *SH3D21* was negatively correlated with DNAsi of HCC cells **(Fig 7H)**, but there was no significant correlation between *SH3D21* and RNAsi. Immunotherapy data in GEO database was further analyzed to observe the intervention effect of immunotherapy on *SH3D21*. These data included Melanoma Treatment study GSE78220, Renal Cancer Treatment Study GSE67501, Urothelial Cancer Treatment Study IMvigor, and immunotherapy data from TCIA database (https://tcia.at/patients). It was found that none of the above immunotherapies had a statistically significant effect on *SH3D21* (*P* >0.05), thus indicating that *SH3D21* was not sensitive to immunotherapy. Thereafter, cancer-related drug sensitivity genomics (GDSC, Genomics of Drug Sensitivity in Cancer)were used to screen *SH3D21* sensitive drugs. The top 10 sensitive drugs identified were AMG-706, ATRA, CEP-701, GSK1904529A, JNK-9L, JQ12, LFM-A13, PFI-1, SB505124 and YK4-279.

### Alterations in *SH3D21* during HCC cell development, construction of predictive models and *SH3D21* expression in pan-cancer

A mimetic time-series analysis was performed to observe the potential changes in *SH3D21* during the development of HCC cells. The results revealed that *SH3D21* was at a relatively stable level during the development of HCC cells and did not show any significant decrease or increase with the progression of HCC **(Fig 9A, B)**.

**Fig. 9.**
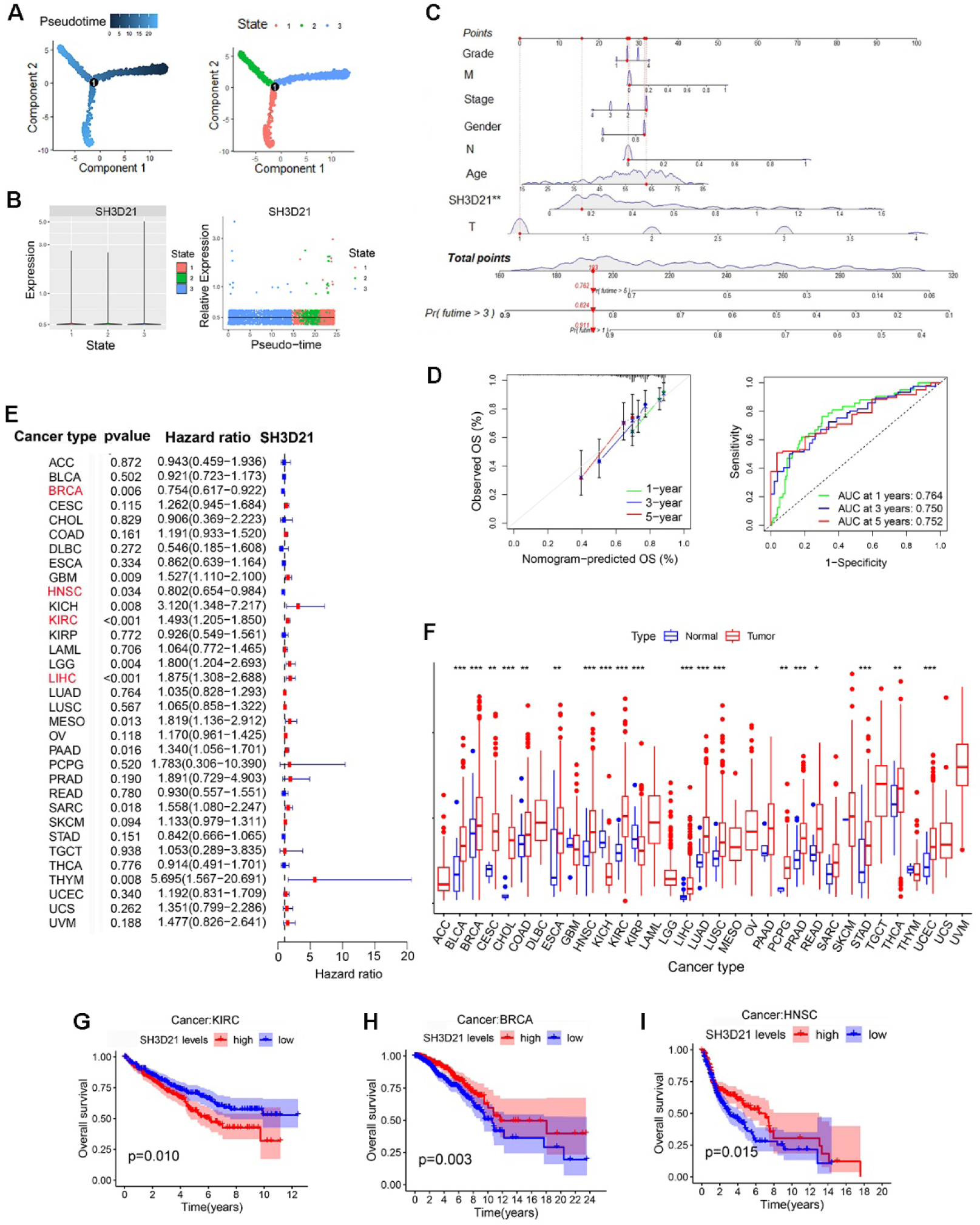
Alterations of *SH3D21* during HCC development, construction of predictive models and SH3D21 expression in pan-cancer. **A, B** Proposed temporal analysis of *SH3D21* in development of HCC. **C** Nomograph of prognostic factors predicting OS of patients based on multiple variables (including: *SH3D21* expression, patient age, gender, hepatocellular carcinoma stage, grade). **D** ROC curve construction of expression level of *SH3D21* and its predictive value for survival of HCC. **E** Univariate analysis of *SH3D21* on the prognosis of 33 tumor patients. **F** Differential analysis of *SH3D21* expression in 24 tumors. **G-I** Effect of high *SH3D21* expression on the prognosis of patients with KIRC (G), BRCA (H) and HNSC (I).

To aid clinicians assess patients’ prognoses based on the aforementioned parameters, we established a nomograph of different prognostic factors including *SH3D21* expression, patient age, gender, hepatocellular carcinoma stage, grade, and several other variables to predict the OS of the patients **(Fig 9C).** The multivariate ROC curve analysis demonstrated that *SH3D21* expression possessed predictive value for one-year (AUC=0.764), three-year (AUC=0.750) and five-year (AUC=0.752) survival rates in HCC patients **(Fig 9D)**, the fitting curve revealed that the model had good reliability.

Finally, the mRNA expression and clinical data of *SH3D21* in 33 tumors were downloaded from the UCSC Xena online database (https://xena.ucsc.edu/) to observe the differential expression of *SH3D21* in different tumors and the effect on patient prognosis. We set the number of normal sample cases in the tumors to be greater than or equal to 1. A total of 24 tumors were included in the analysis. Interestingly, the differential expression of *SH3D21* was observed in 19 tumors in comparison to the normal tissues. The expression of *SH3D21* was elevated in 17 tumors (*P*<0.05), decreased in 2 tumors (*P*<0.05), and did not differ in the remaining 5 tumors (*P*>0.05) **(Fig 9F)**. We divided *SH3D21* into two groups, high and low, according to the median value of *SH3D21* expression in 19 tumors, and observed the effect of different expression on patients’ prognosis. High *SH3D21* expression could lead to poor prognosis in patients with HCC in addition to kidney renal clear cell carcinoma (KIRC) **(Fig 9G)**, whereas high *SH3D21* expression could prolong patients’ survival in head and neck squamous cell carcinoma (HNSC) **(Fig 9I)** and breast invasive carcinoma (BRCA) **(Fig 9H)**. The results of a further univariate analysis indicated that *SH3D21* could be used as an influential factor affecting patient prognosis in LIHC, KIRC, BRCA and HNSC tumor types **(Fig 9E)**. This result suggested that the function of *SH3D21* in various tumors varies depending on the tumor type.

## 4. Discussion

In this study, the role of *SH3D21* in HCC was investigated in detail. The results revealed that expression level of *SH3D21* was increased in HCC and associated with poor prognosis of HCC patients. *SH3D21* was found to promote the proliferation, invasion, and migration of HCC cells, and can activate PI3K/AKT/ mTOR signaling pathway.

*SH3D21* gene is located at 1p34.3 locus and encodes a protein with a molecular weight of 71 kDa. In terms of subcellular localization, *SH3D21* is found in both the nucleus and the cytoplasmic membrane. Functionally, *SH3D21* can function as a signaling molecule within the cells by forming complexes with other proteins. For instance, in a previous study by Jacklyn N. Hellwege et al., it was discovered that two single nucleotide polymorphisms within the *SH3D21* gene were associated with Resting Metabolic Rates (RMR) based on exome array analysis [15]. Moreover, in another study, Mohammad Masoudi et al. utilized the Pickles database and identified the requirement of *SH3D21* gene for maintaining the survival of NCIH526 lung cancer cell line. Furthermore, in the context of pancreatic cancer research, *SH3D21* has been identified as a novel sensitizer to gemcitabine treatment [23]. Despite these findings, our understanding of the exact role played by *SH3D21* in tumorigenesis remains relatively limited. Hence, further research is warranted to comprehensively elucidate the potential involvement of *SH3D21* in HCC. In the analysis of HCC samples from the TCGA database, SH3D21 was found to be highly expressed in HCC, and the increased expression of *SH3D21* was positively correlated with the histological grade, clinical stage, and T stage of HCC, thereby suggesting that *SH3D21* has a key role in promoting the clinical progression of HCC. This conclusion was also confirmed by the analysis of the HCC dataset from the ICGC database. The IOBR package was further employed to analyze the correlation between *SH3D21* and TME characteristics. TCGA and ICGC database analysis consistently revealed that high-expression of *SH3D21* was associated with EMT, cell cycle, DNA replication, TH1, TH2 cell regulation, MDSC and TIP immune cell infiltration. These results indicated that *SH3D21* can effectively promote the clinical progression of HCC by stimulating the proliferation and invasion of HCC cells and generating tumor immunosuppressive microenvironment. To further investigate the cell types that *SH3D21* specifically acts on, HCC single-cell sequencing dataset from the GEO database was employed and by analyzing the expression of *SH3D21* in different types of cells, the primary cellular role of *SH3D21* was determined. The results revealed that *SH3D21* was expressed on HCC cells, macrophages, monocytes, B cells, T cells and NK cells. This indicated that *SH3D21* had important functional effects on all the above cells. In this study, only HCC cells were selected for follow-up research.

A single-cell dataset from the GEO database revealed that *SH3D21* was expressed at higher levels in HCC samples in comparison to the normal hepatocytes. The role of *SH3D21* in promoting the proliferation and invasion of HCC cells was further verified by CCK8, cloning, invasion, and migration experiments. In order to further explore the specific mechanism of *SH3D21* promoting the progression of HCC cells, GSEA analysis results of TCGA database, KEGG analysis results of ICGC database and GSVA analysis results of single cell sequencing dataset were used. The results of enrichment analysis revealed that cell cycle, PI3K-Akt, DNA replication, mismatch repair, gap link, and VEGF angiogenesis pathways were significantly enriched in all the above three datasets. However, the potential effect of *SH3D21* on activation of PI3K-Akt signaling pathway was the primary focus of this study. The PI3K-Akt signaling pathway has been implicated in the regulation of cell proliferation, invasion, apoptosis signaling processes. Activation of PI3K pathway can not only promote the cell cycle progression and tumor cell migration, but also inhibit immune cells, thereby constructing a tumor immunosuppressive microenvironment.

Activation of the PI3K/AKT/mTOR signaling pathway can occur through diverse mechanisms, including AKT-induced phosphorylation of apoptosis and cell cycle blocking factors such as BAD (BCL2-related cell death agonist), MDM2 (ubiquitin ligase of p53), FOXO (apoptosis-inducing transcription factor), IKK (inhibitor of kappa B kinase), and IκB (IκB kinase, which acts as a inhibitor of the transcription factor NF-κB). Interestingly, targeted inhibition of these factors can lead to enhanced activity of cell cycle, DNA replication, and other associated signaling pathways [24, 25]. In addition, due to metabolic reprogramming in tumors, abnormal activation of PI3K/AKT/mTOR pathway can promote the immunosuppressive response [26], which in turn can lead to the formation of the immunosuppressive network of tumors [27]. PI3K/AKT/ mTOR signaling pathway also plays a role in regulating M1/M2 polarization of macrophages [28], similar to the tumor-associated macrophage (TAM) switch between immune stimulation and immune suppression [29]. In addition, PI3K/AKT/mTOR signaling pathway also plays a significant role in promoting glycolytic metabolism in HCC cells, which can cause the abnormal accumulation of lactic acid. As a product of glycolysis, lactic acid produced by the tumor cells can promote M2-like polarization of tumor-associated macrophages [30–32]. Moreover, M2-like polarization of tumor-associated macrophages can inhibit anti-tumor immunity, stimulate tumor angiogenesis, enhance tumor cell invasion as well as infiltration, and further promote the tumor growth [33–35], thus forming a vicious cycle. In this study, the activation of PI3K/AKT/ mTOR signaling pathway by high expression of *SH3D21* was confirmed by further experiments.

Considering that there are few studies reported on potential role of *SH3D21* in tumorigenesis, the expression of *SH3D21* in pan-cancer and its influence on pan-cancer were also analyzed. Among the 33 different tumor types investigated, *SH3D21* exhibited altered expression in 19 tumor tissues in comparison to the normal tissues. Specifically, *SH3D21* expression was increased in 17 tumors (*P*<0.05) and decreased in 2 tumors (*P*<0.05). Interestingly, although *SH3D21* expression was elevated in certain tumors such as BLCA, ESCA, and READ, it did not significantly impact the overall patient survival (*P*>0.05). These observations indicate that the influence of *SH3D21* might significantly vary across different tumor types.

## 5. Conclusion

In summary, *SH3D21* can display elevated expression levels in various tumor types, including primary HCC. Notably, *SH3D21* can promote the proliferation and invasion of HCC by activating PI3K/AKT/mTOR signaling pathway. Consequently, *SH3D21* could serve as a promising candidate for targeted intervention in hepatoma treatment.

## Data Availability

All relevant data are within the manuscript and its Supporting Information files.

## Supporting information

**S1 Raw images** (PDF)

**S2 Raw images** (PDF)

## Author contributions

**Conceptualization:** Ning Luo

**Data curation:** Wangxia Tong, Tao Lu

**Formal analysis:** Li Liu, Jibing Chen

**Methodology:** Rong Liu, Li Liu, Wangxia Tong, Tao Lu

**Resources:** Wangxia Tong, Ning Luo and Jibing Chen

**Supervision:** Jibing Chen

**Validation:** Jibing Chen

**Writing-original draft:** Wangxia Tong

**Writing-review & editing:** Wangxia Tong, Ning Luo

## Acknowledgments

Not applicable

## Consent for publication

This article has been authorized by all authors and agreed to be published.

## Ethical approval

The studies involving human participants were reviewed and approved by the Ethics Committees of Ruikang Hospital Affiliated to Guangxi University of Chinese Medicine. The assigned ethical review approval number: KY2022-045.The atients/participants provided their written informed consent to participate in this study.

## References

1. Villanueva A. Hepatocellular Carcinoma. New Engl J Med. 2019;380(15):1450–62. 10.1056/NEJMra1713263 PMID:30970190

2. Forner A, Reig M, Bruix J. Hepatocellular carcinoma. Lancet. 2018;391(10127):1301-14. 10.1016/S0140-6736(18)30010-2 PMID:29307467

3. Wong KM, King GG, Harris WP. The Treatment Landscape of Advanced Hepatocellular Carcinoma. Curr Oncol Rep. 2022;24(7):917–27. 10.1007/s11912-022-01247-7 PMID:35347594 PMID:35347594

4. Hartke J, Johnson M, Ghabril M. The diagnosis and treatment of hepatocellular carcinoma. Semin Diagn Pathol. 2017;34(2):153–9. 10.1053/j.semdp.2016.12.011 PMID:28108047

5. Kelley RK, Rimassa L, Cheng AL, Kaseb A, Qin S, Zhu AX, et al. Cabozantinib plus atezolizumab versus sorafenib for advanced hepatocellular carcinoma (COSMIC-312): a multicentre, open-label, randomised, phase 3 trial. Lancet Oncol. 2022;23(8):995–1008. 10.1016/S1470-2045(22)00326-6 PMID:35798016

6. Yau T, Park JW, Finn RS, Cheng AL, Mathurin P, Edeline J, et al. Nivolumab versus sorafenib in advanced hepatocellular carcinoma (CheckMate 459): a randomised, multicentre, open-label, phase 3 trial. Lancet Oncol. 2022;23(1):77–90. 10.1016/S1470-2045(21)00604-5 PMID:34914889

7. McGlynn KA, Petrick JL, El-Serag HB. Epidemiology of Hepatocellular Carcinoma. Hepatology. 2021;73 Suppl 1(Suppl 1):4–13. 10.1002/hep.31288 PMID:32319693

8. Manning BD, Toker A. AKT/PKB Signaling: Navigating the Network. Cell. 2017;169(3):381–405. 10.1016/j.cell.2017.04.001 PMID:28431241

9. Sever R, Brugge JS. Signal transduction in cancer. Csh Perspect Med. 2015;5(4). 10.1101/cshperspect.a006098 PMID:25833940

10. Sun EJ, Wankell M, Palamuthusingam P, McFarlane C, Hebbard L. Targeting the PI3K/Akt/mTOR Pathway in Hepatocellular Carcinoma. Biomedicines. 2021;9(11). 10.3390/biomedicines9111639 PMID:34829868

11. Porta C, Paglino C, Mosca A. Targeting PI3K/Akt/mTOR Signaling in Cancer. Front Oncol. 2014;4:64. 10.3389/fonc.2014.00064 PMID:24782981

12. Yu XN, Chen H, Liu TT, Wu J, Zhu JM, Shen XZ. Targeting the mTOR regulatory network in hepatocellular carcinoma: Are we making headway? Bba-Rev Cancer. 2019;1871(2):379–91. 10.1016/j.bbcan.2019.03.001 PMID:30951815

13. Yang C, Zhang H, Zhang L, Zhu AX, Bernards R, Qin W, et al. Evolving therapeutic landscape of advanced hepatocellular carcinoma. Nat Rev Gastro Hepat. 2023;20(4):203–22. 10.1038/s41575-022-00704-9 PMID:36369487

14. Tian LY, Smit DJ, Jucker M. The Role of PI3K/AKT/mTOR Signaling in Hepatocellular Carcinoma Metabolism. Int J Mol Sci. 2023;24(3). 10.3390/ijms24032652 PMID: 36768977

15. Hellwege JN, Velez ED, Acra S, Chen K, Buchowski MS, Edwards TL. Association of gene coding variation and resting metabolic rate in a multi-ethnic sample of children and adults. Bmc Obes. 2017;4:12. 10.1186/s40608-017-0145-5 PMID:28417008

16. Cerami E, Gao J, Dogrusoz U, Gross BE, Sumer SO, Aksoy BA, et al. The cBio cancer genomics portal: an open platform for exploring multidimensional cancer genomics data. Cancer Discov. 2012;2(5):401–4. 10.1158/2159-8290.CD-12-0095 PMID:22588877

17. Goldman MJ, Craft B, Hastie M, Repecka K, McDade F, Kamath A, et al. Visualizing and interpreting cancer genomics data via the Xena platform. Nat Biotechnol. 2020;38(6):675–8. 10.1038/s41587-020-0546-8 PMID:32444850

18. Liao Y, Wang J, Jaehnig EJ, Shi Z, Zhang B. WebGestalt 2019: gene set analysis toolkit with revamped UIs and APIs. Nucleic Acids Res. 2019;47(W1): W199-205. 10.1093/nar/gkz401 PMID:31114916

19. Slovin S, Carissimo A, Panariello F, Grimaldi A, Bouche V, Gambardella G, et al. Single-Cell RNA Sequencing Analysis: A Step-by-Step Overview. Methods Mol Biol. 2021; 2284:343–65. 10.1007/978-1-0716-1307-8_19 PMID:33835452

20. Hu C, Li T, Xu Y, Zhang X, Li F, Bai J, et al. CellMarker 2.0: an updated database of manually curated cell markers in human/mouse and web tools based on scRNA-seq data. Nucleic Acids Res. 2023;51(D1):D870–6. 10.1093/nar/gkac947 PMID:36300619

21. Qiu X, Mao Q, Tang Y, Wang L, Chawla R, Pliner HA, et al. Reversed graph embedding resolves complex single-cell trajectories. Nat Methods. 2017;14(10):979–82. 10.1038/nmeth.4402 PMID:28825705

22. Lu Y, Yang A, Quan C, Pan Y, Zhang H, Li Y, et al. A single-cell atlas of the multicellular ecosystem of primary and metastatic hepatocellular carcinoma. Nat Commun. 2022;13(1):4594. 10.1038/s41467-022-32283-3 PMID:35933472

23. Masoudi M, Seki M, Yazdanparast R, Yachie N, Aburatani H. A genome-scale CRISPR/Cas9 knockout screening reveals SH3D21 as a sensitizer for gemcitabine. Sci Rep-Uk. 2019;9(1):19188. 10.1038/s41598-019-55893-2 PMID:31844142

24. Mossmann D, Park S, Hall MN. mTOR signalling and cellular metabolism are mutual determinants in cancer. Nat Rev Cancer. 2018;18(12):744–57. 10.1038/s41568-018-0074-8 PMID:30425336

25. Giannone G, Ghisoni E, Genta S, Scotto G, Tuninetti V, Turinetto M, et al. Immuno-Metabolism and Microenvironment in Cancer: Key Players for Immunotherapy. Int J Mol Sci. 2020;21(12). 10.3390/ijms21124414 PMID:32575899

26. Okkenhaug K. Signaling by the phosphoinositide 3-kinase family in immune cells. Annu Rev Immunol. 2013;31:675–704. 10.1146/annurev-immunol-032712-095946 PMID:23330955

27. Speiser DE, Ho PC, Verdeil G. Regulatory circuits of T cell function in cancer. Nat Rev Immunol. 2016;16(10):599–611. 10.1038/nri.2016.80 PMID:27526640

28. Tsai CF, Chen GW, Chen YC, Shen CK, Lu DY, Yang LY, et al. Regulatory Effects of Quercetin on M1/M2 Macrophage Polarization and Oxidative/Antioxidative Balance. Nutrients. 2021;14(1). 10.3390/nu14010067 PMID:35010945

29. Zhang Q, Lou Y, Bai XL, Liang TB. Immunometabolism: A novel perspective of liver cancer microenvironment and its influence on tumor progression. World J Gastroentero. 2018;24(31):3500–12. 10.3748/wjg.v24.i31.3500 PMID:30131656

30. Kaneda MM, Messer KS, Ralainirina N, Li H, Leem CJ, Gorjestani S, et al. PI3Kgamma is a molecular switch that controls immune suppression. Nature. 2016;539(7629):437-42. 10.1038/nature19834 PMID:27642729

31. Wu X, Chen H, Wang Y, Gu Y. Akt2 Affects Periodontal Inflammation via Altering the M1/M2 Ratio. J Dent Res. 2020;99(5):577–87. 10.1177/0022034520910127 PMID:32228353

32. Vergadi E, Ieronymaki E, Lyroni K, Vaporidi K, Tsatsanis C. Akt Signaling Pathway in Macrophage Activation and M1/M2 Polarization. J Immunol. 2017;198(3):1006–14. 10.4049/jimmunol.1601515 PMID:28115590

33. Colegio OR, Chu NQ, Szabo AL, Chu T, Rhebergen AM, Jairam V, et al. Functional polarization of tumour-associated macrophages by tumour-derived lactic acid. Nature. 2014;513(7519):559-63. 10.1038/nature13490 PMID:25043024

34. Sunakawa Y, Stintzing S, Cao S, Heinemann V, Cremolini C, Falcone A, et al. Variations in genes regulating tumor-associated macrophages (TAMs) to predict outcomes of bevacizumab-based treatment in patients with metastatic colorectal cancer: results from TRIBE and FIRE3 trials. Ann Oncol. 2015;26(12):2450–6. 10.1093/annonc/mdv474 PMID:26416897

35. Xiang X, Wang J, Lu D, Xu X. Targeting tumor-associated macrophages to synergize tumor immunotherapy. Signal Transduct Tar. 2021;6(1):75. 10.1038/s41392-021-00484-9 PMID: 33619259

